# Epidemiological Investigation of New SARS-CoV-2 Variant of Concern 202012/01 in England

**DOI:** 10.1101/2021.01.14.21249386

**Authors:** Alfredo Maria Gravagnuolo, Layla Faqih, Cara Cronshaw, Jackie Wynn, Lewis Burglin, Paul Klapper, Mark Wigglesworth

## Abstract

Lighthouse Labs network tests for the presence of RNA of SARS-CoV-2, the causative agent of COVID-19. The Thermofisher TaqPath assay targets three regions of SARS-CoV-2; ORF1ab, N and S-genes. The assay identified a drop in S-gene target detection among positive samples due to the circulation of a new SARS-CoV-2 Variant of Concern (VOC) designated as 202012/01. By end of December 2020, 60% of daily positive test results at Alderley Park Lighthouse Labs, were linked to the new Variant of Concern. This timeline view identifies the rapid spread of the variant across the country.

## INTRODUCTION

Alderley Park (AP) Lighthouse Lab (LHL) tests for the presence of RNA of the Severe Acute Respiratory Syndrome Coronavirus 2 (SARS-CoV-2), the causative agent of Coronavirus disease 2019 (COVID-19).

The laboratory uses the ThermoFisher TaqPath™ COVID-19 test, for real-time reverse transcription polymerase chain reaction (RT-qPCR) detection of SARS-CoV-2 in nose and throat swabs. The assay contains three primer/probe sets specific to different SARS-CoV-2 genomic regions, and the RNA of the bacteriophage MS2 is used as an internal control molecule to monitor potential sample inhibition.

As viruses can show substantial genetic variability, which can over time result in mismatches between the primer/probe and the target sequences, the assay targets three genomic sequences (ORF1ab, N and S) to improve reliability of detection of SARS-Cov-2.

An increase in the number of positive samples with lack of S gene amplification was noted in late November. As a percentage of positive samples from the LH laboratories are referred to the Wellcome Sanger Laboratory at Cambridge (UK) for whole genome sequencing we were able to identify that the samples with failure to detect S gene amplification were due to a deletion of six nucleotides in the S-gene, which results in the loss of two amino acids of the Spike protein at positions 69 and 70 (Δ69-70); (PHE, 2020). Data from the national network of high throughput testing facilities has been utilised to demonstrate the localisation and rapid temporal spread of this variant. Samples for SARS-CoV-2 detection are derived from any area of the UK. A distribution network load balances sample distribution to one of the five major UK centres on a daily basis, analysis of samples arriving in one of these facilities can provide information on the distribution of infection within the whole of England.

## RESULTS

### The impact of the new variant on the diagnostic testing

The proportion in England of positive specimens tested using the ThermoFisher TaqPath™ COVID-19 assay with failure of S-gene target detection increased rapidly in December 2020 rising to over 70% of strong positive test results detected within the facility at the beginning of January 2021 (***Figure 1***, five-day rolling rate). The failure of S-gene detection does not significantly alter the clinical validity of the test result as detection of the alternate target (ORF1Ab and N) has remained robust.

**Figure 1.**
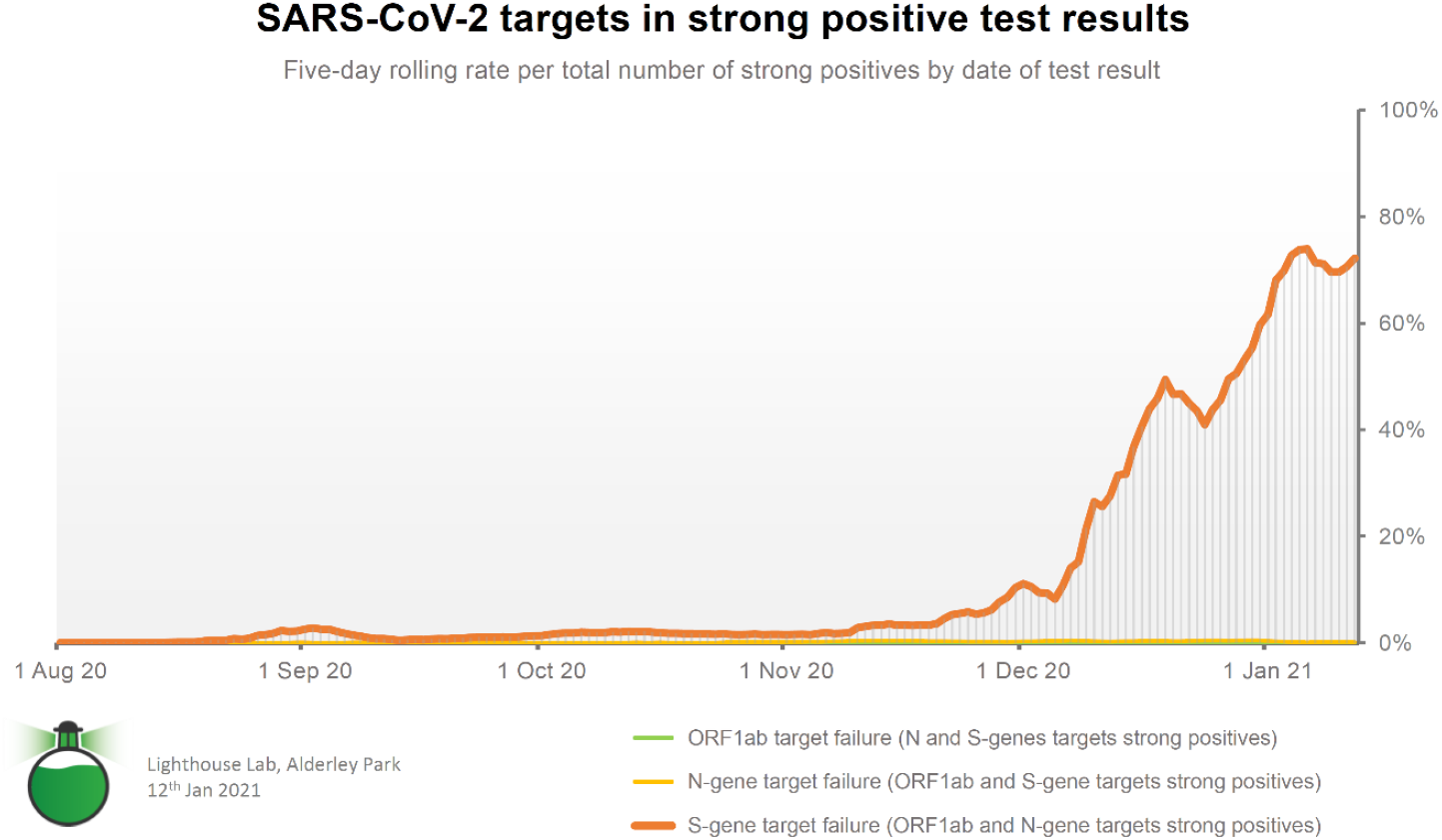
TaqPath™ COVID 19 Assay, S-gene detection in comparison to ORF1ab and N-gene.

### Epidemiological clusters associated with S-gene target failure

Samples tested at Alderley Park Lighthouse Labs, from the 1^st^ to the 21^st^ of December 2020, were collected and linked to geographical area within England (***Table 1***; ***Figure 2***; ***supporting information video map***).

**Table 1.** Geographical origins in England of the new cases of S-gene target failure at Alderley Park Lighthouse Labs, in the assessed time ranges.

| Region – Data from 1<sup>st</sup> - 21<sup>st</sup> Dec | [%] | Region – Data from 1<sup>st</sup> - 30<sup>th</sup> Dec | [%] |
| --- | --- | --- | --- |
| London | 38.8 | London | 27.6 |
| South East | 30.1 | South East | 24.3 |
| North West | 9.9 | North West | 22.5 |
| East of England | 7.1 | East of England | 5.1 |
| South West | 6.7 | South West | 9.9 |
| West Midlands | 4.6 | West Midlands | 7.7 |
| East Midlands | 2.1 | East Midlands | 1.6 |
| Yorkshire and the Humber | 0.7 | Yorkshire and the Humber | 1.2 |
| North East | 0.1 | North East | 0.1 |
| **Total** | **100** | **Total** | **100** |

**Figure 2.**
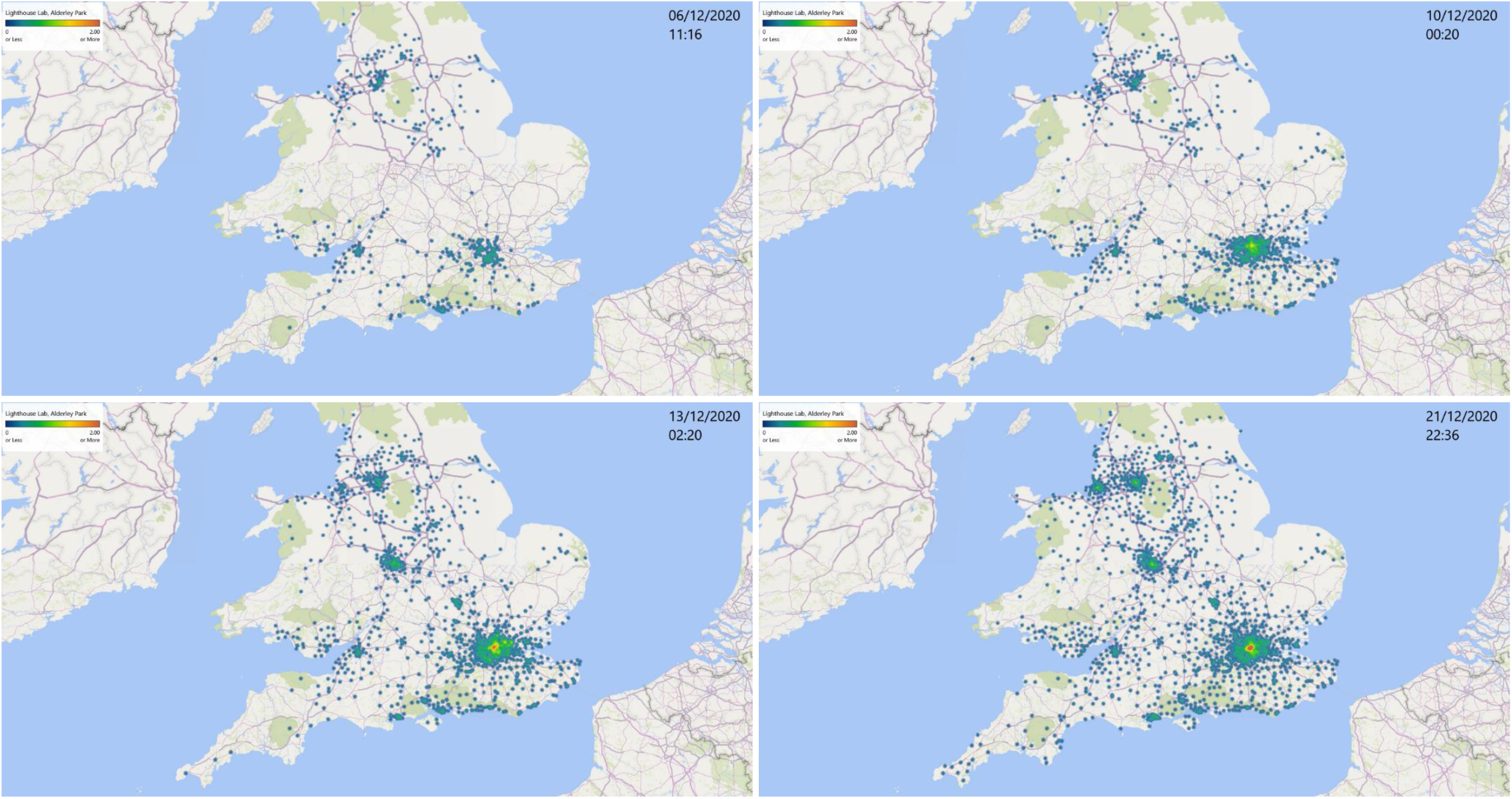
Map of S-gene detection failure, associated with the new SARS-CoV-2 variant. Data collected at Alderley Park, Lighthouse Labs, from 1^st^ to 21^st^ of December 2020.

The distribution of S-gene target failure cases shows a relatively higher burden in London, East-South East, parts of the North West, South west regions and West Midlands. While the sample collection and distribution network load balancing does not allow that the same number of samples is delivered consistently from each geographic area the data does provide some indication on the national geographic spread of the variant with time. As sample numbers were not collected equally from each region a low detection rate cannot be considered conclusive proof of the variant not being present. However, when viewed as a percentage of samples the data clearly shows a geographical bias within the overall sample set. Clustering was observed around population centres of London, cities of the coastal South, Manchester, Birmingham, Bristol and Liverpool (***Figure 2***).

Areas with the highest incidence in our study also coincide with areas now reporting high levels of patient hospitalisation (NHS, 2021). The timeline view identifies the rapid spread of this in high population areas across the country and reinforces the necessity of subsequent National lockdown to contain spread of the variant.

### Molecular analysis of Virus strains associated with S-gene target failure – Pending Work

Samples of extracted RNA tested at Alderley Park, Lighthouse Labs, were sequenced at the Wellcome Sanger Institute, Cambridge, UK.

At the beginning of December, the Sanger Institute has identified and sequenced a significant number of samples with negative S-gene target detection by the TaqPath™ COVID 19 Kit, but positive for the ORF1ab and N-gene targets. Analysis showed that Spike protein mutations characteristic of VOC 202012/01 were present in these S-gene target detection failure cases indicating the predictive nature of our S-gene target detection failure for the presence of the VOC. However, ongoing molecular surveillance is needed to ensure identify mutations, which are currently preventing amplification of the S-gene target, remain characteristic of the VOC.

## CONCLUSIONS

The failure of S-gene target detection is related to the recent emergence of the new variant of concern. Geographical regions affected by *VOC* 202012/01 are clearly illustrated by the absence of S gene detection in the TaqPath assay used in our study, which coincide with areas now reporting high levels of patient hospitalisation.

Ongoing molecular monitoring will be needed to ensure that no further new S gene variants are co-emerging with *VOC* 202012/01.

The value of high throughput testing and use of multitarget PCR technology in early detection of the emergence of viral variants is illustrated in the present data.

## EXPERIMENTAL SECTION

### Sample acquisition

Samples delivered to the LH laboratory, Alderley Park, can be derived from any area of the UK. A distribution network load balances sample distribution to one of the five major UK centres daily. Sample tracking data is held centrally in a proprietary data base (Edge; Department of Health and Social Care, UK).

### SARS-CoV-2 Testing

Nose and throat swab samples in virus transport medium collected at a variety of test centres and home self-collected samples shipped to Alderley Park were extracted using the MagMAX™ Viral/Path-ogen Nucleic Acid Isolation Kit (Thermofisher, Warrington, UK) and Kingfisher Flex extraction platform (ThermoFisher). PCR amplification used the TaqPath™ COVID-19 Combo Kit (Thermofisher) and 384 well format on Quantstudio Flex 7 System and UgenTec-FastFinder PCR analysis software (Eugentec, Hasselt, Belgium). S-gene target detection failure was defined where: ORF1ab Cq value lower than 31.6; N-gene Cq lower than 30.1; S-gene target was not amplified. Those cases were linked to the new SARS-CoV-2 variant.

### Geographical origins of samples

Subject postal district, (***Figure 2*** and ***supporting information video map***), and test region (***Table 1***), were extracted via interrogation of the Edge data.

### Data analysis

***Figure 1***. December data available in ***Table 3*** (last column) showing daily figures and five-day rolling rate of S-gene Target Failure per total number of strong positive test results. The same method was used for the five-day rolling rate the ORF1ab and N-gene target failure per total number of strong positive test results.

**Table 2.** Cq cut-off values for strong positive test results in this study.

| Conditions for strong positive test results<br>at least two target amplicons |  |  |
| --- | --- | --- |
| ORF1ab<br>Cq < 31.6 | N gene<br>Cq < 30.1 | S gene<br>Cq < 31.6 |

**Table 3.**
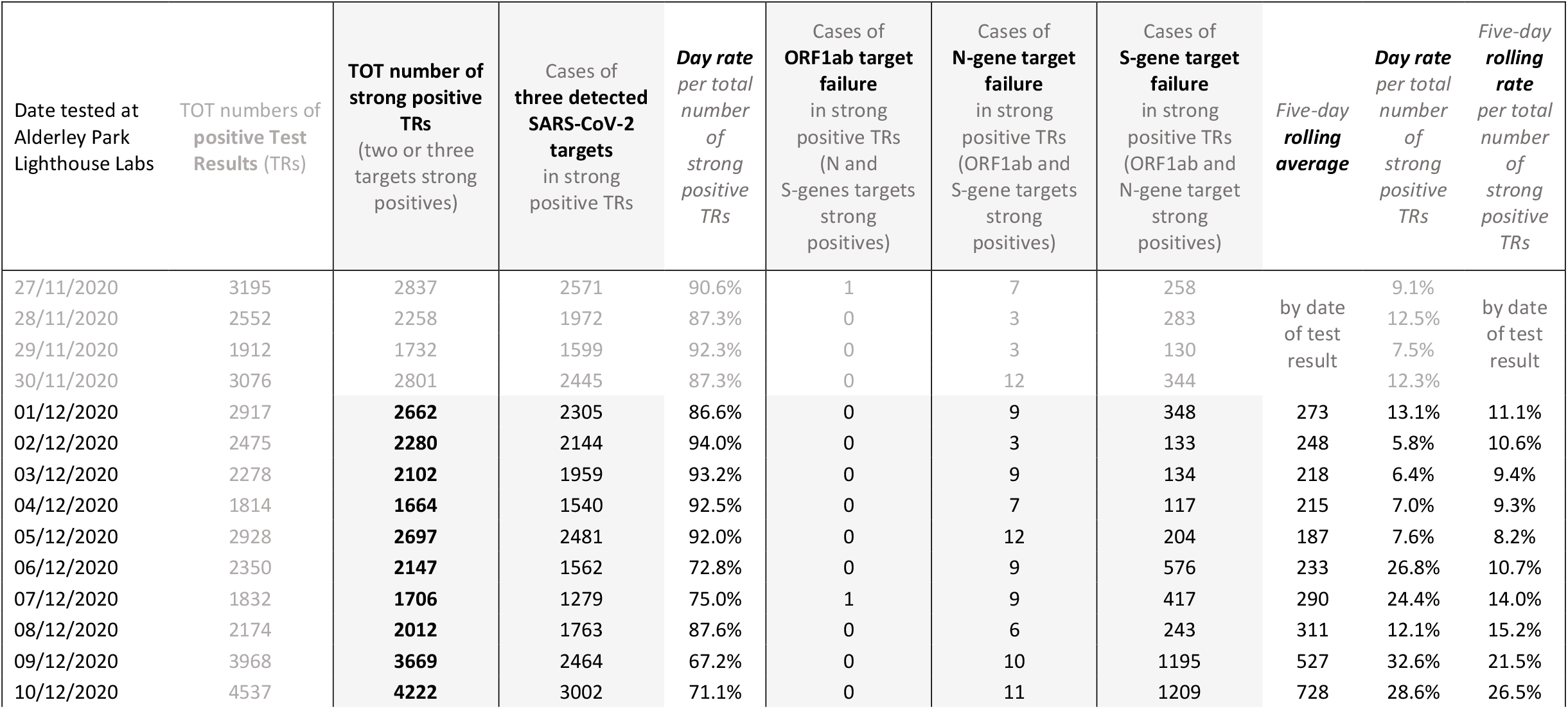
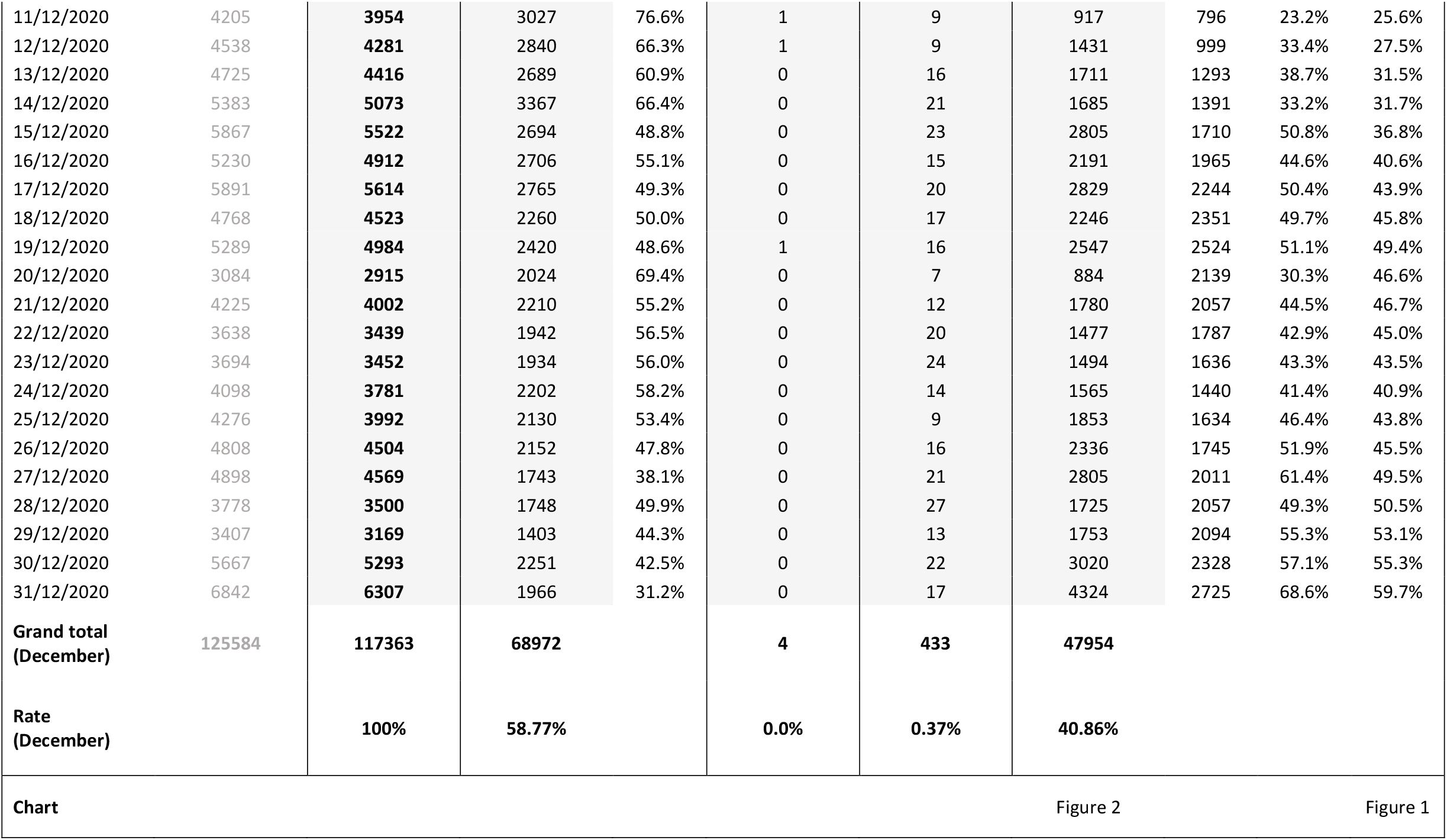
Raw data of December, Alderley Park, Lighthouse Labs.

***Figure 2*** and ***supporting information video map***. Locations displayed on the Map resulted from subject postal district and date tested. Cases displayed on the Map were accumulated over the assessed period in chronological order generating a heat map. Data were limited to numbers (counts) of cases from Alderley Park data set, and not normalised by the total number of positive test results. *Heat map:* indicative density of cases of S-gene target failure in the areas highlighted by colours (see colour bar). *Map coverage:* 98% of cases listed in ***Table 3*** under column “cases of S-gene target failure” were matched with locations in the assessed time range. *Accuracy:* 95% of matched locations where plotted with high confidence. As a result of map coverage and accuracy, ∼93% of total cases in ***Table 3*** were linked to their locations and indicated on the heat map.

***Table 1***. Geographical origins of new cases of S-gene target failure, identified at Alderley Park Lighthouse Labs from 1^st^ – 21^st^ December 2020, were shown as fractions of total cases of S-gene target failure in the same time range. For comparison, a longer assessment period, 1^st^ - 30^th^ December, was shown.

## Supporting information

supporting information video map

## Data Availability

Full data available upon formal request addressed to the corresponding author Layla Faqih for auditing purposes

https://lighthouselabsalderleypark.org.uk/

## ACKNOWLEDGMENTS

*COVID-19 National Testing Programme*

Health Research Authority, National Health Service (**HRA-NHS**), United Kingdom.

Medicines Discovery Catapult, Lighthouse Labs, Alderley Park (**MDC-LHL-AP**), United Kingdom: Simon Chapman, Andrzej Rutkowski, Helen Hind, Salina Hussain, Nathan Hayes, Thomas Ingram, Farah Tahmasebi, Sul Mulroy, Maged Taema, Kirsten Hobby, Amir Mossanen-Parsi.

The University of Manchester, United Kingdom (**UOM**). Department of Health & Social Care, United Kingdom (**DHSC**). Wellcome Sanger Institute, United Kingdom (**WSI**): Jeffrey Barrett. Astra Zeneca, United Kingdom (**AZ**): David Murray.

## WEB

https://lighthouselabsalderleypark.org.uk/

Published: January 2021.

Version 1, release date 07/01/2021 (submitted).

Version 1.1, release date 14/01/2021 (submitted).

