## Supplementary figures and images for "Epidemiological Investigation of New SARS-CoV-2 Variant of Concern 202012/01 in England"

### supporting information video map

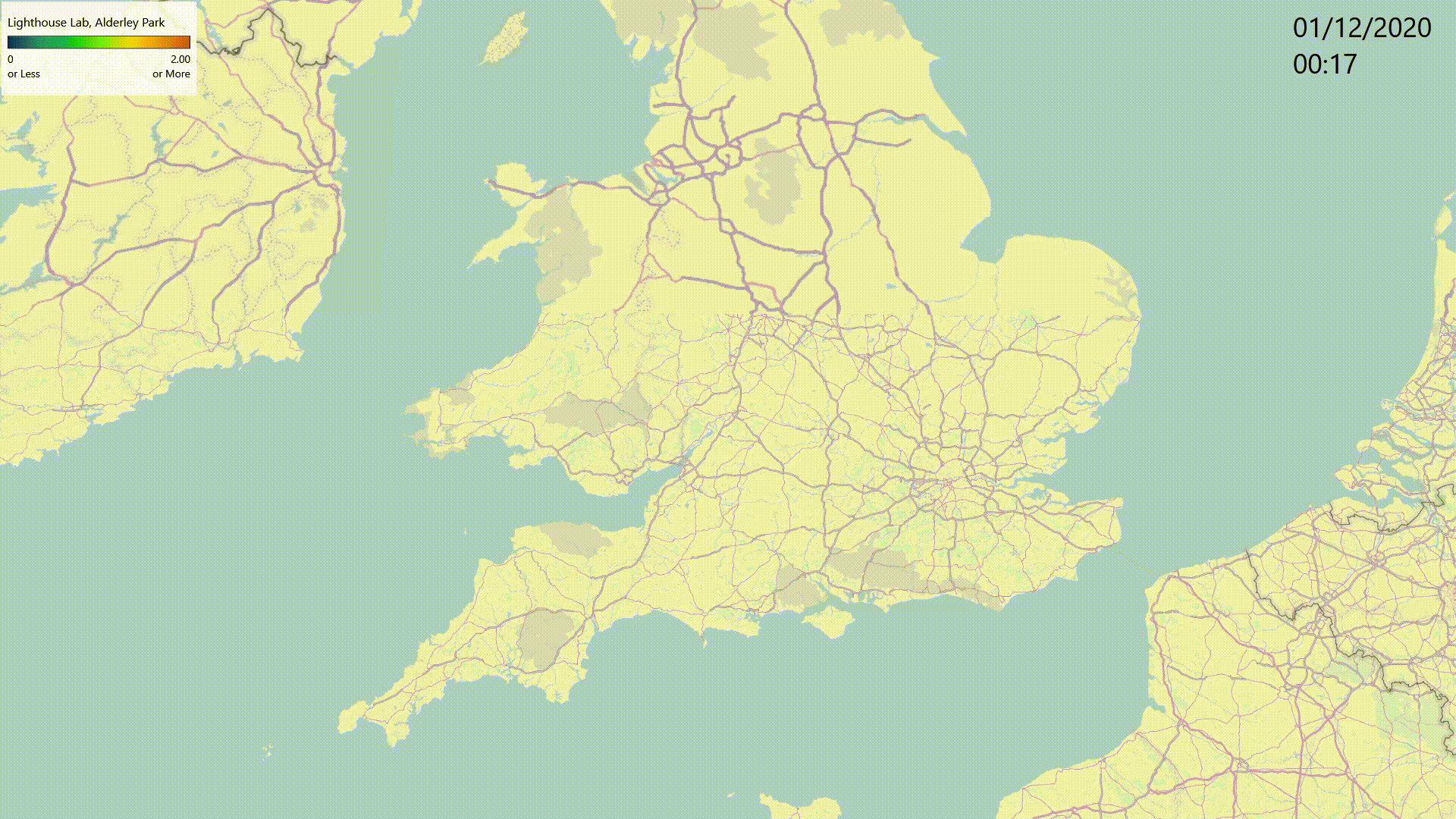
